# Educational attainment and mental health conditions: a within-sibship Mendelian randomization study

**DOI:** 10.1101/2024.08.10.24311789

**Authors:** María Fernanda Vinueza Veloz, Laxmi Bhatta, Paul Remy Jones, Martin Tesli, George Davey Smith, Neil Martin Davies, Ben M. Brumpton, Øyvind Erik Næss

## Abstract

**Importance:** Observational studies have demonstrated consistent protective effects of higher educational attainment (EA) on the risk of suffering mental health conditions (MHC). Determining whether these beneficial effects are causal is challenging given the potential role of dynastic effects and demographic factors (assortative mating and population structure) in this association.

**Objective:** To evaluate to what extent the relationship between EA and various MHC is independent from dynastic effects and demographic factors.

**Design:** Within-sibship Mendelian randomization (MR) study.

**Setting:** One-sample MR analyses included participants’ data from the Trøndelag Health Study (HUNT, Norway) and UK Biobank (United Kingdom). For two-sample MR analyses we used summary statistics from publicly available genome-wide-association-studies.

**Participants:** 61 880 siblings (27 507 sibships).

**Exposure:** Years of education.

**Main outcomes:** Scores for symptoms of anxiety, depression and neuroticism using the Hospital Anxiety Depression Scale (HADS), the 7-item Generalized Anxiety Disorder Scale (GAD-7), the 9-item Patient Health Questionnaire (PHQ-9), and the Eysenck Personality Questionnaire, as well as self-reported consumption of psychotropic medication.

**Results:** One standard deviation (SD) unit increase in years of education was associated with a lower symptom score of anxiety (−0.20 SD [95%CI: −0.26, −0.14]), depression (−0.11 SD [−0.43, 0.22]), neuroticism (−0.30 SD [−0.53, −0.06]), and lower odds of psychotropic medication consumption (OR: 0.60 [0.52, 0.69]). Estimates from the within-sibship MR analyses showed some attenuation, which however were suggestive of a causal association (anxiety: −0.17 SD [−0.33, −0.00]; depression: −0.18 SD [-1.26, 0.89]; neuroticism: −0.29 SD [−0.43, −0.15]); psychotropic medication consumption: OR, 0.52 [0.34, 0.82]).

**Conclusions and Relevance:** Associations between EA and MHC in adulthood, although to some extend explained by dynastic effects and demographic factors, overall remain robust, indicative of a causal effect. However, larger studies are warranted to improve statistical power and further validate our conclusions.

**Key points:** *Question:* Are the protective effects of higher educational attainment on the risk of suffering mental health conditions independent from familial (i.e.,dynastic effects) and demographic factors (i.e., assortative mating and population structure)?

*Findings:* Although to some extend explained by familial and demographic factors, overall the protective effects of higher educational attainment remain robust.

*Meaning:* Our findings are consistent with those of previous observational studies, which suggests that higher educational attainment may causally reduce the risk of suffering a mental health condition.

## Introduction

Mental health conditions (MHC) are mental disorders, psycho-social disabilities as well as mental states associated with significant distress, impaired functioning, or risk of self-harm ^1^. Common MHC including anxiety and mood disorders (e.g., depression) are among the top causes of years lived with disability across all age groups ^2^. Moreover, people with MHC have a mortality rate that is at least twice higher than that of the general population or people without such conditions ^3^. Socioeconomic circumstances including those related to level of educational attainment (EA) have been related to the risk of suffering MHC ^4^.

Higher EA is associated with a lower risk of suffering MHC including, anxiety, depression, personality disorders, and substance abuse ^5–7^. This inverse relationship might be explained by the wider availability of mental/physical resources in adulthood to cope with daily hassles among educated individuals, which could assist preventing MHC ^8^. Nevertheless, it has been shown that family-related factors influence both educational level and mental health status, which suggests that alternative causal pathways in adulthood may have a role ^9–12^.

Mendelian randomization (MR) is a method based on instrumental variable analysis used to investigate relationships between exposures and outcomes by using genetic variants or polygenic scores (PGS) as instruments for the modifiable factor ^13^. MR relies on the premise that there is a causal pathway from an individual’s genotype to the individual’s phenotype (e.g., a genotype-phenotype association). If its three core assumptions hold, the approach can overcome both exposure-outcome confounding and reverse causation.

To provide evidence of causality i) the genetic instrument must be associated to the exposure of interest and this relationship is required to be reasonably strong (relevance); ii) there mustn’t be common factors associated to the genetic instrument or the outcome of interest (independence); iii) the genetic instrument must influence the outcome only through the exposure of interest, which implies that the effect of the genetic instrument on an outcome is fully mediated by the exposure (exclusion) ^13^.

Two recent population-based MR studies have provided evidence that supports the role of EA in the aetiology of MHC like anxiety and depression in adulthood ^14,15^. However, since the phenotypic variation of complex traits cannot be fully attributed to direct genetic effects, such MR estimates might be biased ^16,17^. In fact, violations of the independence assumption resulting from indirect genetic effects (i.e., dynastic effects) and confounding due demographic factors (i.e., assortative mating and population stratification) are now well documented for various traits including education ^16–18^.

Dynastic effects are the result of genetic nurturing, namely the effect of parents’ or other relatives’ genotypes on an individual’s phenotype through their contributions to the environment ^17,18^. Assortative mating is when partners are not selected at random but based on particular characteristics ^19^. Population structure is the presence of subpopulation differences in allele frequencies that correlate with both the phenotype and the genotype ^16^. In the context of MR all three mechanisms have the potential of amplifying the effect of a given genotype-phenotype association ^17,20^.

The objective of the present study is to evaluate if the association between EA and symptoms of anxiety, depression, and neuroticism as well as the consumption of psychotropic medication are explained by indirect genetic effects and/or are confounded by demographic factors. We applied a within-sibship design coupled with genomic data to perform MR to account for dynastic effects, assortative mating, and population structure.

### Methodology

#### Study design and data sources

The present is the report of a within-sibship MR study. One-sample and two-sample MR methods were applied using individual-level data, and summary statistics from genome wide association studies (GWAS). Individual-level data came from the Trøndelag Health Study (HUNT) and UK Biobank (UKB) ^21,22^. A brief description of contributing GWAS can be found in eTable 1.

#### Setting and participants

HUNT is a population-based cohort study that is held in the Trøndelag County in Norway and started in 1984 ^21^. We used data from all participants of the second (HUNT2) and third HUNT wave (HUNT3) who had been genotyped. From those, we selected all individuals who were > 30 years of age when they participated in the survey, and had at least one sibling. The final sample from HUNT2 and HUNT3 included 26 770 (10 428 sibships) and 16 718 siblings (7010 sibships), respectively (eTable 2).

The UKB is a prospective cohort study that began in 2006. UKB is following nearly 500 000, 40 to 69 years old participants from across the UK, who volunteered to be part of the study and provided consent for follow-up through linkage to their health records ^22^. We included all participants who had been genotyped. After restricting the sample to sibships with two or more individuals, our analysis sample included 35 118 participants from 17 079 sibships (eTable 2).

#### Genetic variants

For HUNT participants we used a weighted PGS as an instrumental variable for EA (PGS-edu). The calculation of the PGS-edu was based on the genetic variants reported as significantly associated to years of education at the genome-wide level (*p* < 5 × 10^-8^) in a recent GWAS ^23^. From the 3952 independent genetic variants reported by Okbay et al., we included those that were well imputed in the target population (eTable 3) ^24^. For further information see eMethods.

#### Exposure and outcomes

EA was the exposure of interest and was assessed trough the question: What is your highest level of education? for HUNT participants, or Which of the following qualification do you have? for UKB participants. A number of years of attained education was assigned for each of the answers based on The International Standard Classification of Education (ISCED) mapping 1997 (eTable 4).

Symptoms of anxiety, depression and neuroticism as well as consumption of psychotropic medication were the outcomes of interest. In HUNT symptoms of anxiety and depression were assessed by the Hospital Anxiety and Depression Scale (HADS), and in UKB by the 7-item Generalized Anxiety Disorder Scale (GAD-7) and 9-item Patient Health Questionnaire (PHQ-9) ^25–27^. In both HUNT and UKB, neuroticism was measured using the Eysenck Personality Questionnaire and consumption of psychotropic medication was self-reported ^28^. A detailed description on how the exposure and outcomes were processed can be found in eMethods.

#### Ethics approval and informed consent

See eMethods.

#### Statistical analysis

Before running the analyses using individual-level data we standardized all numerical variables so that they had a mean of 0 and standard deviation (SD) of 1. EA, as well as symptoms of anxiety, depression and neuroticism were analysed as continuos, while consumption of psychotropic medication as categorical (yes/no). The association between EA and the outcomes was analysed by applying one-sample MR (two-stage least squares regression), and ordinary least squares (OLS) or logistic (LOG) regression for comparison.

All models were adjusted by sex and age. However, when the PGS-edu was included as predictor, the model was also adjusted by the first 10 principal components of ancestry (PCA) and genotyping batch to account for population structure. In all cases we assumed that standard errors were correlated within sibships and therefore clustered standard errors were computed. OLS, LOG and one-sample MR analyses were performed using the “feols” (for continuos) and “felgm” (for categorical) functions of the “fixest” package in R ^29,30^.

Any difference between families due to indirect genetic effects (i.e. dynastic effects) or bias due to assortative mating was accounted for by using family fixed effects (anxiety, depression and neuroticism) or the sibling difference method (consumption of psychotropic medication) ^16^. All analyses were performed using the “fixest” package in R (see eMethods for a detailed description)^30^.

Individual-level data for each of the two cohorts were analysed separately using the same model specifications and R packages. Then, results were meta-analysed using the “rma” function from the “metafor” R package ^31^. We applied fixed effect models, except when heterogeneity between HUNT and UK Biobank estimates was detected, that is Cochran’s Q Chi^2^ < 0.05 and I^2^ > 50% (eTable 5).

Two-sample MR analyses were performed using summary statistics from GWAS described in eTable 1 and the R package “TwoSampleMR” ^32^. The inverse-variance estimator weighted (IVW) estimator and its 95% confidence intervals (CI) are reported in the present work. Robust estimators including weighted median, weighted mode, and MR-Egger coefficients were calculated to investigate pleiotropy and reported in supplementary tables. The directionality of the effect was evaluated using Steiger test of directionality. MR-Egger intercept test was performed to assess pleiotropy. Within-sibship two-sample MR was performed for depressive symptoms and neuroticism, as only for them we had access to summary statistics from within-sibship GWAS (eTable 1) ^33^.

#### Handling of missing information

We imputed missing data for various questions of the HADS score as well as education and consumption of psychotropic medication in HUNT (see eTable 6 for details on missing information). We applied multivariate imputation to each HUNT survey, using fully conditional specification implemented by the MICE algorithm ^34^. For each imputed data set, we then calculated the corresponding score. For further information see eMethods.

## Results

### Descriptive statistics

In both cohorts participation rate among females was higher than that among males. Participants from HUNT were younger, but had similar level of education than those from the UKB (Table 1). Symptoms of anxiety were more often reported by HUNT than from UKB participants (8% vs. 3%). About 5% and 9% of HUNT and UKB participants reported symptoms of depression and consumption of psychotropic medication, respectively (Table 1). However, the consumption of psychotropic medication was slightly higher among HUNT than UKB participants. Differences between females and males can be seen in Table 1. For each of the cohorts, the total number of siblings varied depending on the outcome (eTable 2).

**Table 1.**
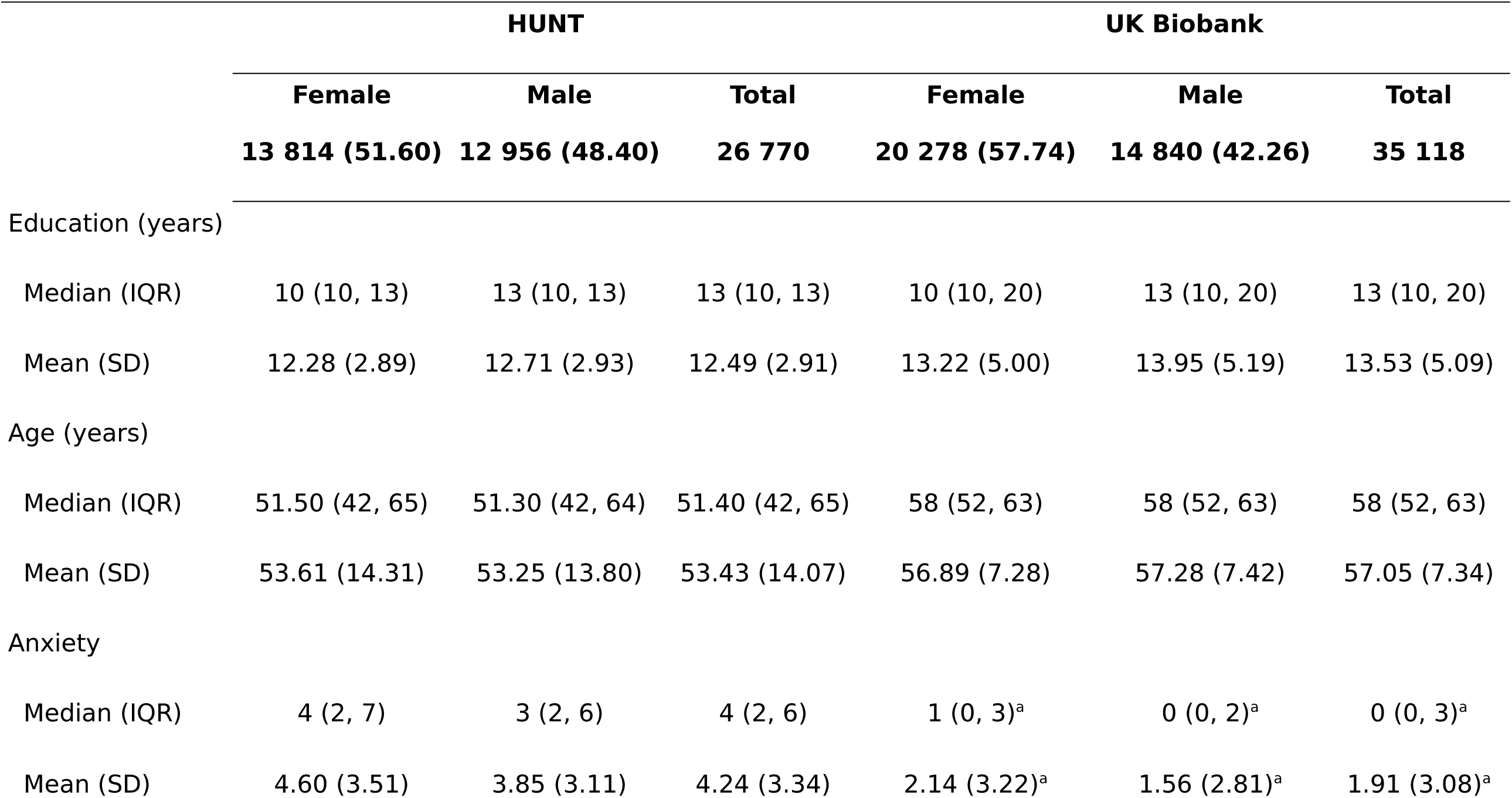
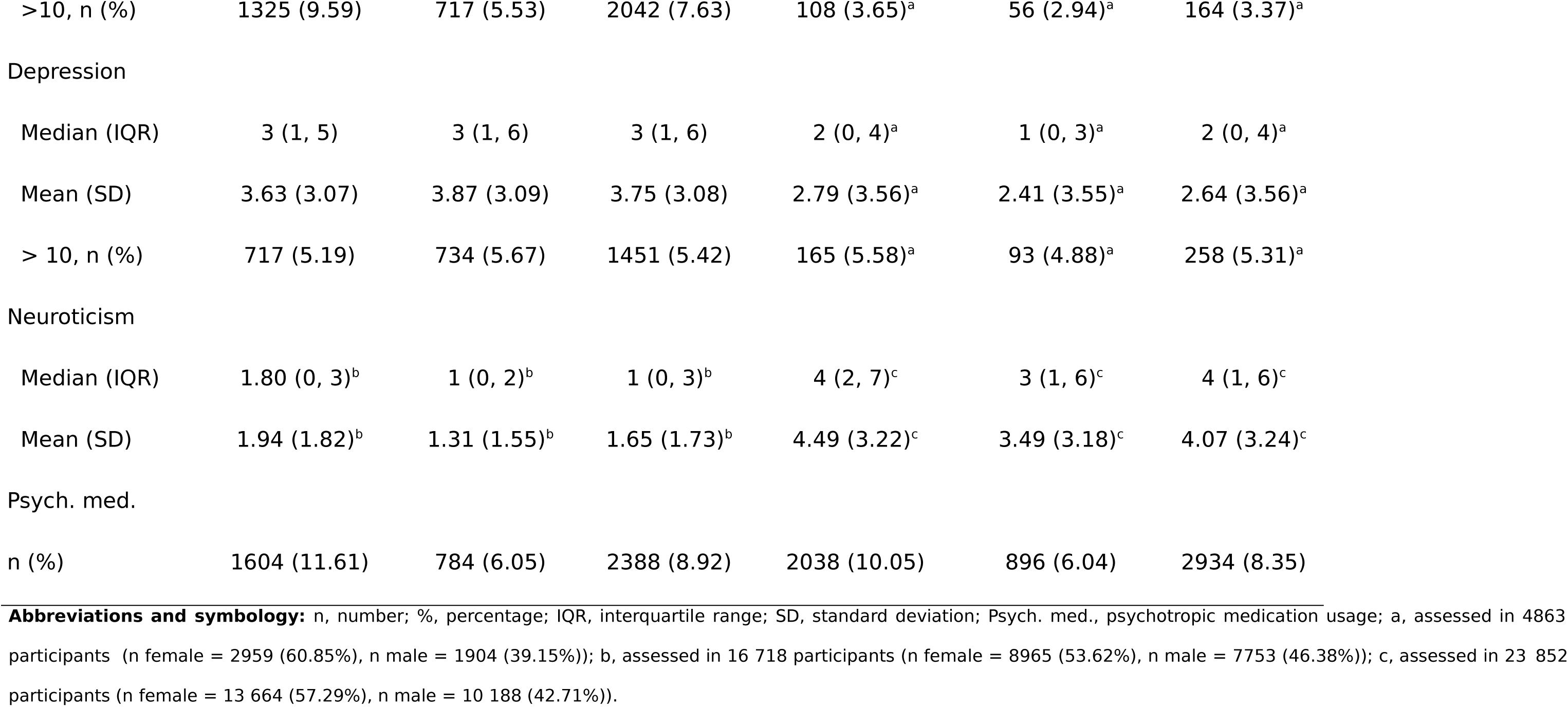
General characteristics of HUNT and UKB samples. . In HUNT symptoms of anxiety and depression were assessed by the Hospital Anxiety and Depression Scale (HADS), and in UK Biobank by the 7-item Generalized Anxiety Disorder Scale (GAD-7) and 9-item Patient Health Questionnaire (PHQ-9), respectively. Neuroticism was assessed using a six-item and a 12-item Eysenck Personality Questionnaire in HUNT and UK Biobank, respectively (see eMethods).

|  | HUNT |  |  | UK Biobank |  |  |
| --- | --- | --- | --- | --- | --- | --- |
|  | Female | Male | Total | Female | Male | Total |
|  | 13 814 (51.60) | 12 956 (48.40) | 26 770 | 20 278 (57.74) | 14 840 (42.26) | 35 118 |
| Education (years) |  |  |  |  |  |  |
| Median (IQR) | 10 (10, 13) | 13 (10, 13) | 13 (10, 13) | 10 (10, 20) | 13 (10, 20) | 13 (10, 20) |
| Mean (SD) | 12.28 (2.89) | 12.71 (2.93) | 12.49 (2.91) | 13.22 (5.00) | 13.95 (5.19) | 13.53 (5.09) |
| Age (years) |  |  |  |  |  |  |
| Median (IQR) | 51.50 (42, 65) | 51.30 (42, 64) | 51.40 (42, 65) | 58 (52, 63) | 58 (52, 63) | 58 (52, 63) |
| Mean (SD) | 53.61 (14.31) | 53.25 (13.80) | 53.43 (14.07) | 56.89 (7.28) | 57.28 (7.42) | 57.05 (7.34) |
| Anxiety |  |  |  |  |  |  |
| Median (IQR) | 4 (2, 7) | 3 (2, 6) | 4 (2, 6) | 1 (0, 3)<sup>a</sup> | 0 (0, 2)<sup>a</sup> | 0 (0, 3)<sup>a</sup> |
| Mean (SD) | 4.60 (3.51) | 3.85 (3.11) | 4.24 (3.34) | 2.14 (3.22)<sup>a</sup> | 1.56 (2.81)<sup>a</sup> | 1.91 (3.08)<sup>a</sup> |
| >10, n (%) | 1325 (9.59) | 717 (5.53) | 2042 (7.63) | 108 (3.65)<sup>a</sup> | 56 (2.94)<sup>a</sup> | 164 (3.37)<sup>a</sup> |
| Depression |  |  |  |  |  |  |
| Median (IQR) | 3 (1, 5) | 3 (1, 6) | 3 (1, 6) | 2 (0, 4)<sup>a</sup> | 1 (0, 3)<sup>a</sup> | 2 (0, 4)<sup>a</sup> |
| Mean (SD) | 3.63 (3.07) | 3.87 (3.09) | 3.75 (3.08) | 2.79 (3.56)<sup>a</sup> | 2.41 (3.55)<sup>a</sup> | 2.64 (3.56)<sup>a</sup> |
| > 10, n (%) | 717 (5.19) | 734 (5.67) | 1451 (5.42) | 165 (5.58)<sup>a</sup> | 93 (4.88)<sup>a</sup> | 258 (5.31)<sup>a</sup> |
| Neuroticism |  |  |  |  |  |  |
| Median (IQR) | 1.80 (0, 3)<sup>b</sup> | 1 (0, 2)<sup>b</sup> | 1 (0, 3)<sup>b</sup> | 4 (2, 7)<sup>c</sup> | 3 (1, 6)<sup>c</sup> | 4 (1, 6)<sup>c</sup> |
| Mean (SD) | 1.94 (1.82)<sup>b</sup> | 1.31 (1.55)<sup>b</sup> | 1.65 (1.73)<sup>b</sup> | 4.49 (3.22)<sup>c</sup> | 3.49 (3.18)<sup>c</sup> | 4.07 (3.24)<sup>c</sup> |
| Psych. med. |  |  |  |  |  |  |
| n (%) | 1604 (11.61) | 784 (6.05) | 2388 (8.92) | 2038 (10.05) | 896 (6.04) | 2934 (8.35) |

**Abbreviations and symbology:** n, number; %, percentage; IQR, interquartile range; SD, standard deviation; Psych. med., psychotropic medication usage; a, assessed in 4863 participants (n female = 2959 (60.85%), n male = 1904 (39.15%)); b, assessed in 16 718 participants (n female = 8965 (53.62%), n male = 7753 (46.38%)); c, assessed in 23 852 participants (n female = 13 664 (57.29%), n male = 10 188 (42.71%)).

The PGS-edu was associated with years of education, conditional from age, sex, first 10 PCA, and batch in both HUNT and UK Biobank. In HUNT, each SD unit increase of the PGS-edu was associated with a 0.19 SD increase in years of education (_95%_CI, 0.18: 0.20, *p =* 4.40×10^-187^, *F-test stat.* = 162.99, *r^2^* = 0.03). In UK Biobank, each SD unit increase of the PGS-edu was associated with a 0.24 SD increase in years of education (_95%_CI, 0.23: 0.24, *p* = 2.20×10^-16^, *F-test stat.* = 103.90, *r^2^ =* 0.06). This association was attenuated after including a family fixed effect (HUNT: 0.13, _95%_CI: 0.11 – 0.15, *p =* 3.47×10^-44^, *F-test stat.* = 1349.62, *r^2^* = 0.64; UK Biobank: 0.13, _95%_CI, 0.11: 0.14, *p* = 3.38×10^-43^, *F-test stat.* = 2362.40, *r^2^* = 0.33). The associations between the PGS ant the outcomes are depicted in eTable 7.

### Main analyses

The direction of the regression and MR estimates was consistent among both cohorts and across all analyses. There were however some differences in the strength of the associations between HUNT and UKB (Table 2). These differences were more evident for the depression and neuroticism MR estimates.

**Table 2.**
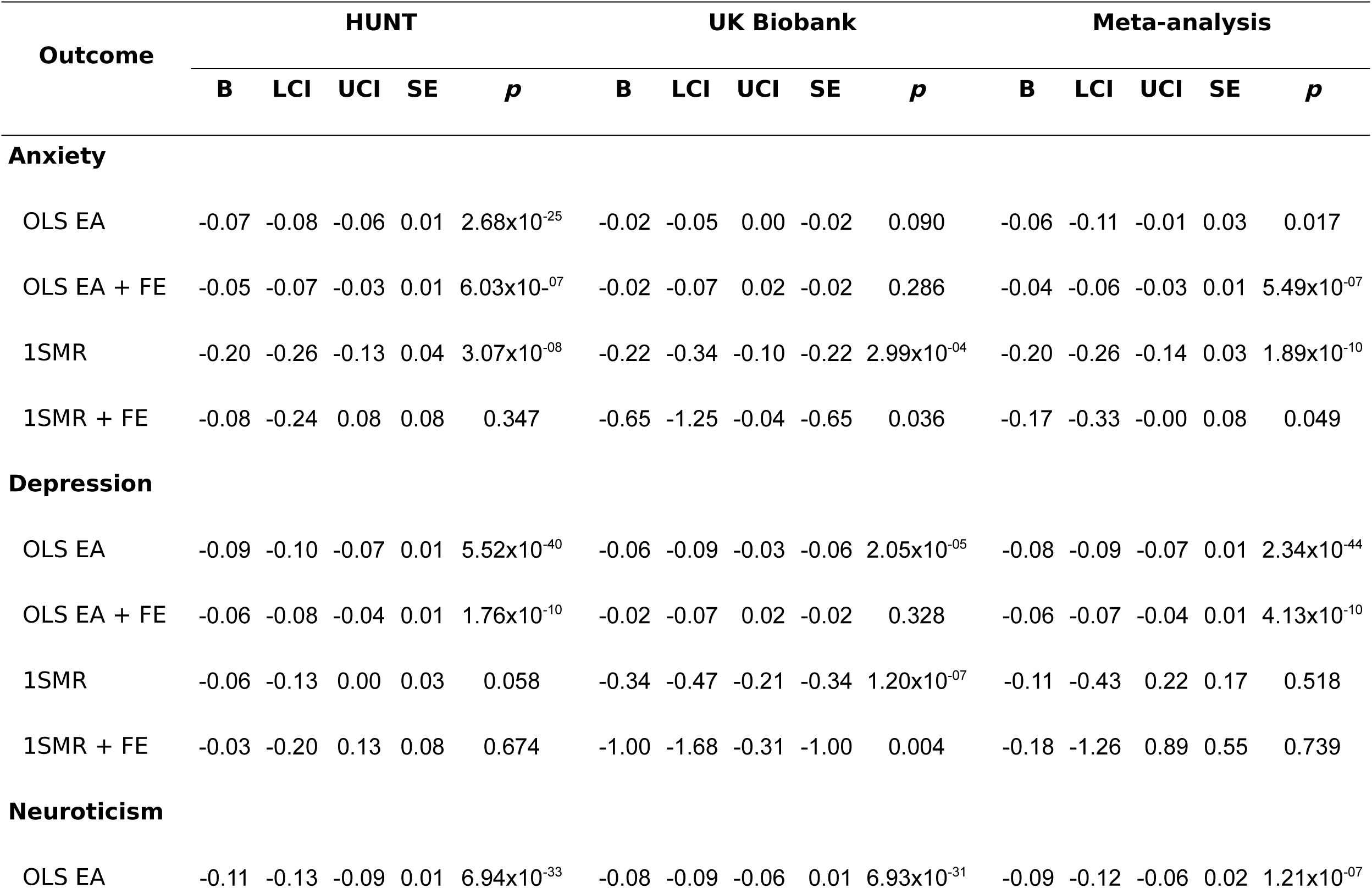
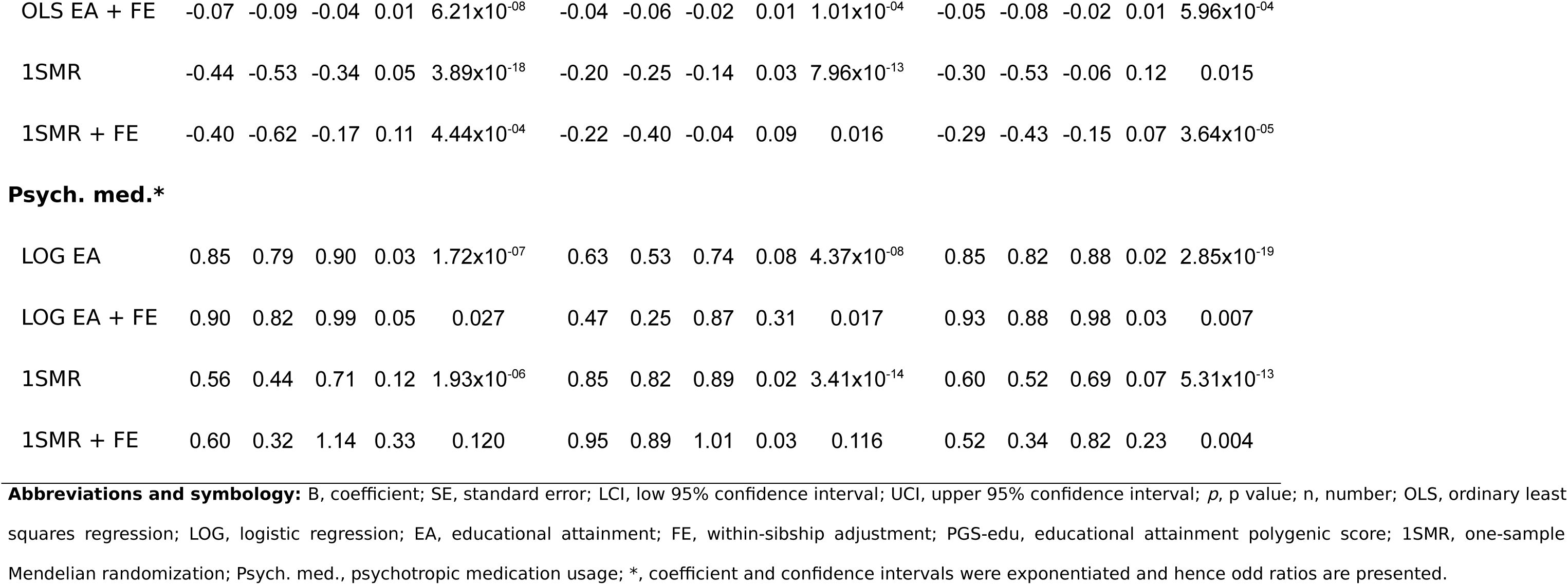
Association between educational attainment and mental health outcomes. . HUNT and UK Biobank data were analysed separately and results then meta-analysed (see Methodology).

| Outcome | HUNT |  |  |  |  | UK Biobank |  |  |  |  | Meta-analysis |  |  |  |  |
| --- | --- | --- | --- | --- | --- | --- | --- | --- | --- | --- | --- | --- | --- | --- | --- |
|  | B | LCI | UCI | SE | *p* | B | LCI | UCI | SE | *p* | B | LCI | UCI | SE | *p* |
| **Anxiety** |  |  |  |  |  |  |  |  |  |  |  |  |  |  |  |
| OLS EA | -0.07 | -0.08 | -0.06 | 0.01 | 2.68x10<sup>-25</sup> | -0.02 | -0.05 | 0.00 | -0.02 | 0.090 | -0.06 | -0.11 | -0.01 | 0.03 | 0.017 |
| OLS EA + FE | -0.05 | -0.07 | -0.03 | 0.01 | 6.03x10<sup>-07</sup> | -0.02 | -0.07 | 0.02 | -0.02 | 0.286 | -0.04 | -0.06 | -0.03 | 0.01 | 5.49x10<sup>-07</sup> |
| 1SMR | -0.20 | -0.26 | -0.13 | 0.04 | 3.07x10<sup>-08</sup> | -0.22 | -0.34 | -0.10 | -0.22 | 2.99x10<sup>-04</sup> | -0.20 | -0.26 | -0.14 | 0.03 | 1.89x10<sup>-10</sup> |
| 1SMR + FE | -0.08 | -0.24 | 0.08 | 0.08 | 0.347 | -0.65 | -1.25 | -0.04 | -0.65 | 0.036 | -0.17 | -0.33 | -0.00 | 0.08 | 0.049 |
| **Depression** |  |  |  |  |  |  |  |  |  |  |  |  |  |  |  |
| OLS EA | -0.09 | -0.10 | -0.07 | 0.01 | 5.52x10<sup>-40</sup> | -0.06 | -0.09 | -0.03 | -0.06 | 2.05x10<sup>-05</sup> | -0.08 | -0.09 | -0.07 | 0.01 | 2.34x10<sup>-44</sup> |
| OLS EA + FE | -0.06 | -0.08 | -0.04 | 0.01 | 1.76x10<sup>-10</sup> | -0.02 | -0.07 | 0.02 | -0.02 | 0.328 | -0.06 | -0.07 | -0.04 | 0.01 | 4.13x10<sup>-10</sup> |
| 1SMR | -0.06 | -0.13 | 0.00 | 0.03 | 0.058 | -0.34 | -0.47 | -0.21 | -0.34 | 1.20x10<sup>-07</sup> | -0.11 | -0.43 | 0.22 | 0.17 | 0.518 |
| 1SMR + FE | -0.03 | -0.20 | 0.13 | 0.08 | 0.674 | -1.00 | -1.68 | -0.31 | -1.00 | 0.004 | -0.18 | -1.26 | 0.89 | 0.55 | 0.739 |
| **Neuroticism** |  |  |  |  |  |  |  |  |  |  |  |  |  |  |  |
| OLS EA | -0.11 | -0.13 | -0.09 | 0.01 | 6.94x10<sup>-33</sup> | -0.08 | -0.09 | -0.06 | 0.01 | 6.93x10<sup>-31</sup> | -0.09 | -0.12 | -0.06 | 0.02 | 1.21x10<sup>-07</sup> |
| OLS EA + FE | -0.07 | -0.09 | -0.04 | 0.01 | 6.21x10<sup>-08</sup> | -0.04 | -0.06 | -0.02 | 0.01 | 1.01x10<sup>-04</sup> | -0.05 | -0.08 | -0.02 | 0.01 | 5.96x10<sup>-04</sup> |
| 1SMR | -0.44 | -0.53 | -0.34 | 0.05 | 3.89x10<sup>-18</sup> | -0.20 | -0.25 | -0.14 | 0.03 | 7.96x10<sup>-13</sup> | -0.30 | -0.53 | -0.06 | 0.12 | 0.015 |
| 1SMR + FE | -0.40 | -0.62 | -0.17 | 0.11 | 4.44x10<sup>-04</sup> | -0.22 | -0.40 | -0.04 | 0.09 | 0.016 | -0.29 | -0.43 | -0.15 | 0.07 | 3.64x10<sup>-05</sup> |
| Psych. med.\* |  |  |  |  |  |  |  |  |  |  |  |  |  |  |  |
| LOG EA | 0.85 | 0.79 | 0.90 | 0.03 | 1.72x10<sup>-07</sup> | 0.63 | 0.53 | 0.74 | 0.08 | 4.37x10<sup>-08</sup> | 0.85 | 0.82 | 0.88 | 0.02 | 2.85x10<sup>-19</sup> |
| LOG EA + FE | 0.90 | 0.82 | 0.99 | 0.05 | 0.027 | 0.47 | 0.25 | 0.87 | 0.31 | 0.017 | 0.93 | 0.88 | 0.98 | 0.03 | 0.007 |
| 1SMR | 0.56 | 0.44 | 0.71 | 0.12 | 1.93x10<sup>-06</sup> | 0.85 | 0.82 | 0.89 | 0.02 | 3.41x10<sup>-14</sup> | 0.60 | 0.52 | 0.69 | 0.07 | 5.31x10<sup>-13</sup> |
| 1SMR + FE | 0.60 | 0.32 | 1.14 | 0.33 | 0.120 | 0.95 | 0.89 | 1.01 | 0.03 | 0.116 | 0.52 | 0.34 | 0.82 | 0.23 | 0.004 |

**Abbreviations and symbology:** B, coefficient; SE, standard error; LCI, low 95% confidence interval; UCI, upper 95% confidence interval; *p*, *p* value; n, number; OLS, ordinary least squares regression; LOG, logistic regression; EA, educational attainment; FE, within-sibship adjustment; PGS-edu, educational attainment polygenic score; 1SMR, one-sample Mendelian randomization; Psych. med., psychotropic medication usage; \*, coefficient and confidence intervals were exponentiated and hence odd ratios are presented.

The results of the MR analyses suggest that EA has a beneficial effect on anxiety, depression and neuroticism symptomatology (Figure 1, 3 and 4). A higher EA was also associated with lower consumption of psychotropic medication (Table 2). A SD unit increase in years of education was associated with lower scores of anxiety (−0.20 SD [−0.26, −0.14]), and neuroticism (−0.30 SD [−0.53, −0.06]) as well as lower odds of psychotropic medication consumption (OR: 0.60 [0.52, 0.69]). There was no association between EA and depression (−0.11 SD [−0.43, 0.22]) (Figure 2). Estimates from one-sample MR were larger than those of the regression analyses, except from depression, but were consistent in direction between each other (Table 2).

**Figure 1.**
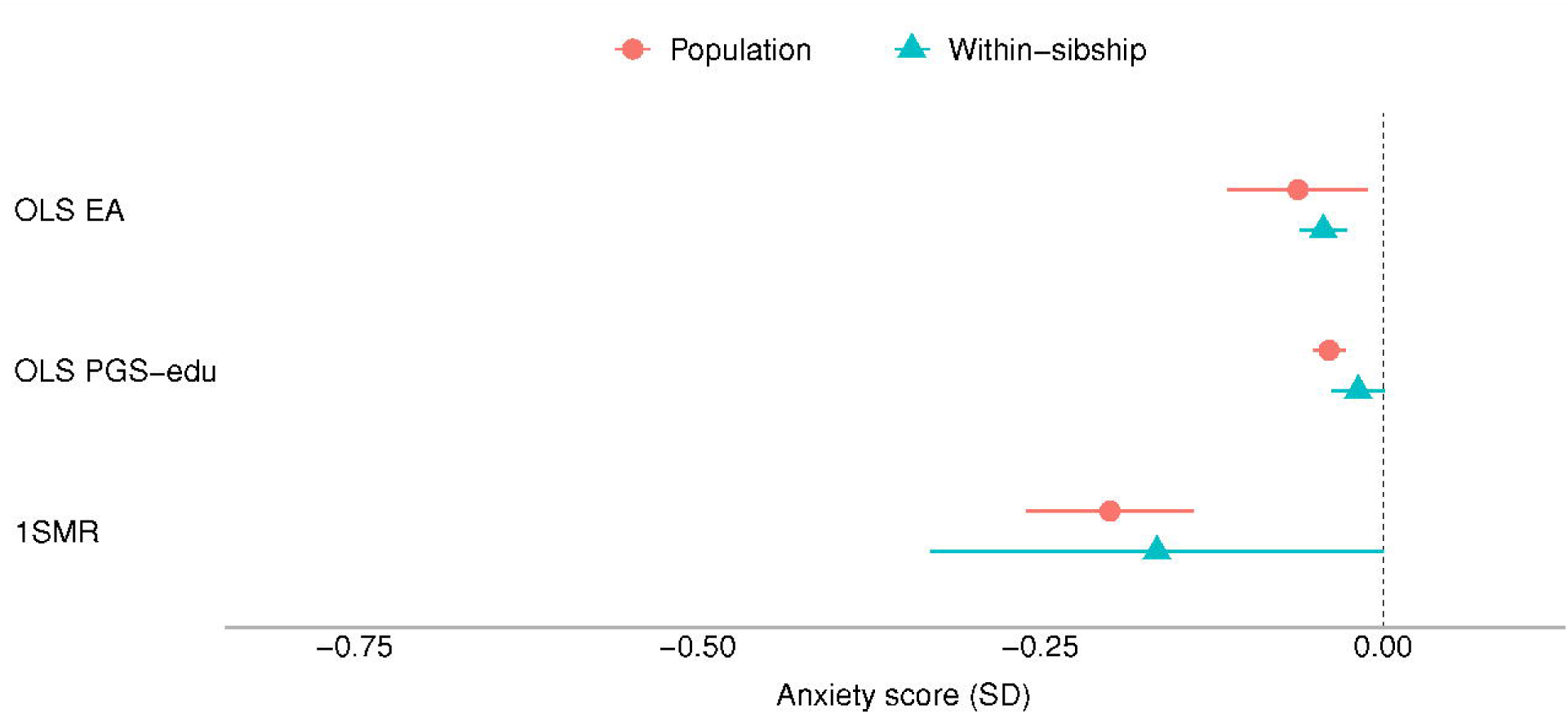
Educational attainment and symptoms of anxiety. Standard deviation (SD) changes in the anxiety score and its 95% confidence interval per SD increase in years of education are shown. Estimated associations are displayed for ordinary least square regression (OLS) and Mendelian randomization models. *Abbreviations: SD, standard deviation unit; OLS EA, ordinary least square regression model with educational attainment as exposure; OLS PGS-edu, ordinary least square regression model with the educational attainment polygenic score as exposure; 1SMR, one-sample Mendelian randomization*.

**Figure 2.**
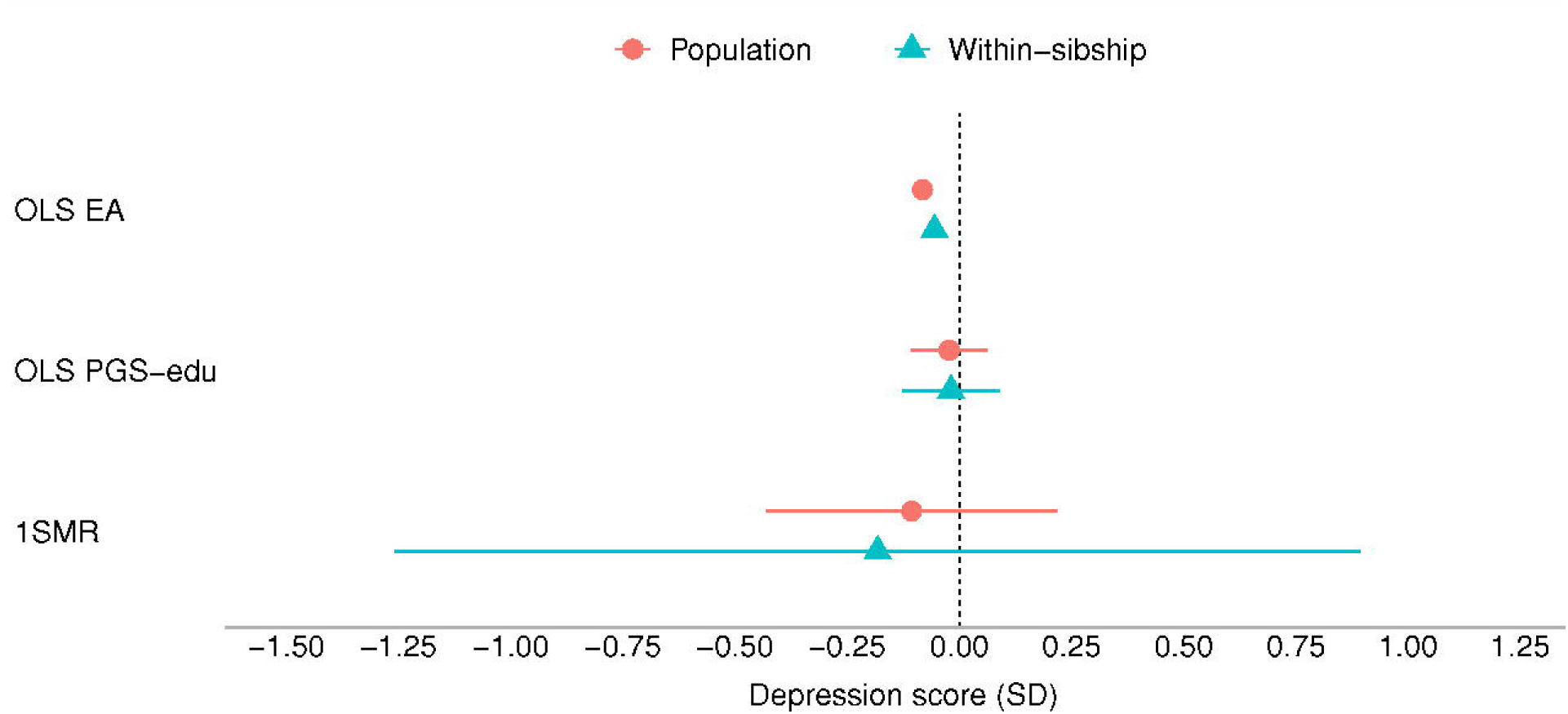
Educational attainment and symptoms of depression. Standard deviation (SD) changes in the depression score and its 95% confidence interval per SD increase in years of education are shown. Estimated associations are displayed for ordinary least square regression (OLS) and Mendelian randomization models. *Abbreviations: SD, standard deviation unit; OLS EA, ordinary least square regression model with educational attainment as exposure; OLS PGS-edu, ordinary least square regression model with the educational attainment polygenic score as exposure; 1SMR, one-sample Mendelian randomization; 2SMR, two-sample Mendelian randomization*.

**Figure 3.**
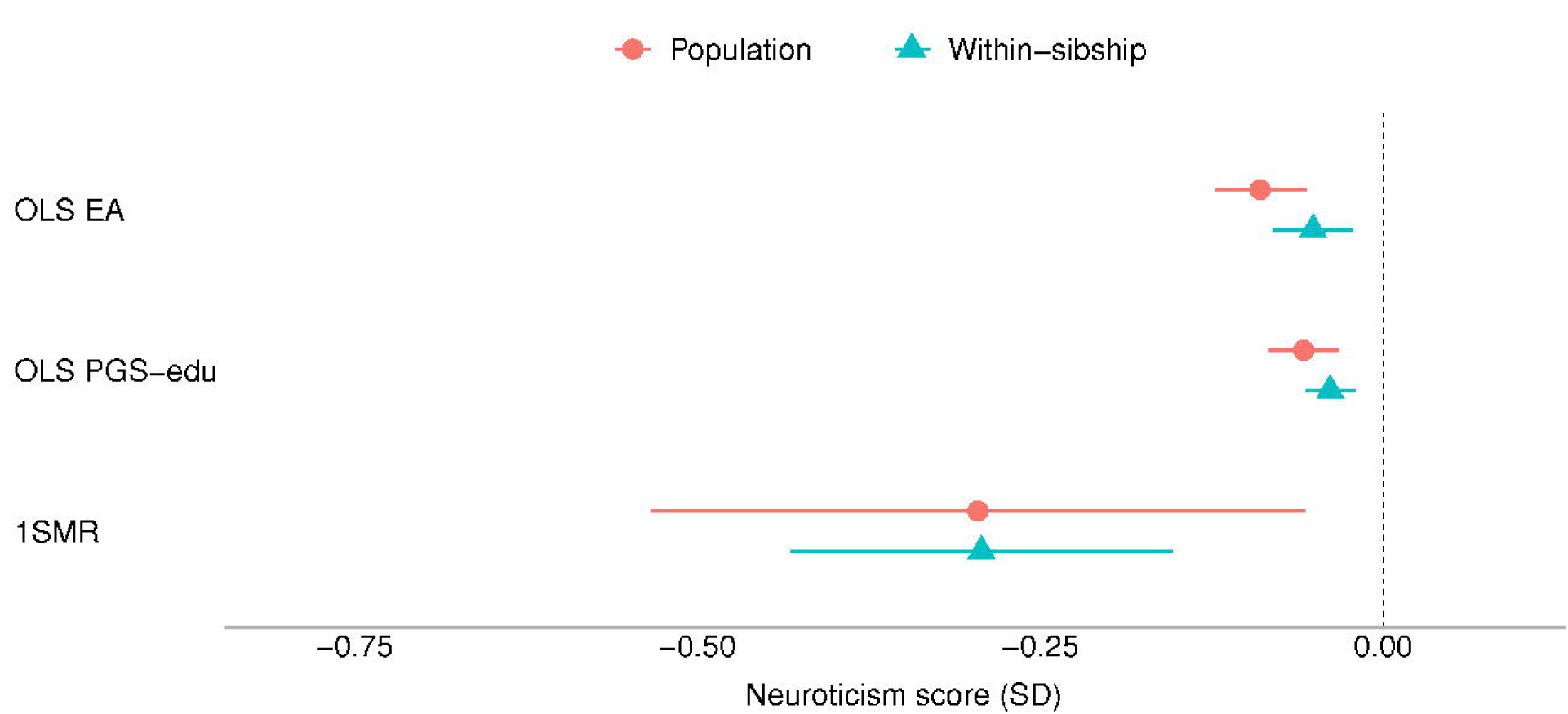
Educational attainment and neuroticism. Standard deviation (SD) changes in the neuroticism score and its 95% confidence interval per SD increase in years of education are shown. Estimated associations are displayed for ordinary least square regression (OLS) and Mendelian randomization models. *Abbreviations: SD, standard deviation unit; OLS EA, ordinary least square regression model with educational attainment as exposure; OLS PGS-edu, ordinary least square regression model with the educational attainment polygenic score as exposure; 1SMR, one-sample Mendelian randomization; 2SMR, two-sample Mendelian randomization*.

Most of the estimates of the within-sibling MR analyses showed some attenuation in comparison to the MR estimates, the exception was depression (Table 2 and Figure 1 to 4). The effect of EA attenuated from −0.20 to −0.17 [−0.33, −0.00] for anxiety, and from −0.30 to −0.29 [−0.43, −0.15] for neuroticism. The odds ration of psychotropic medication decreased from −0.60 to −0.52 [0.33, 0.82]. Some attenuation was also observed among the estimates of the within-sibling regression analyses, except from consumption of psychotropic medication (Table 2).

**Figure 4.**
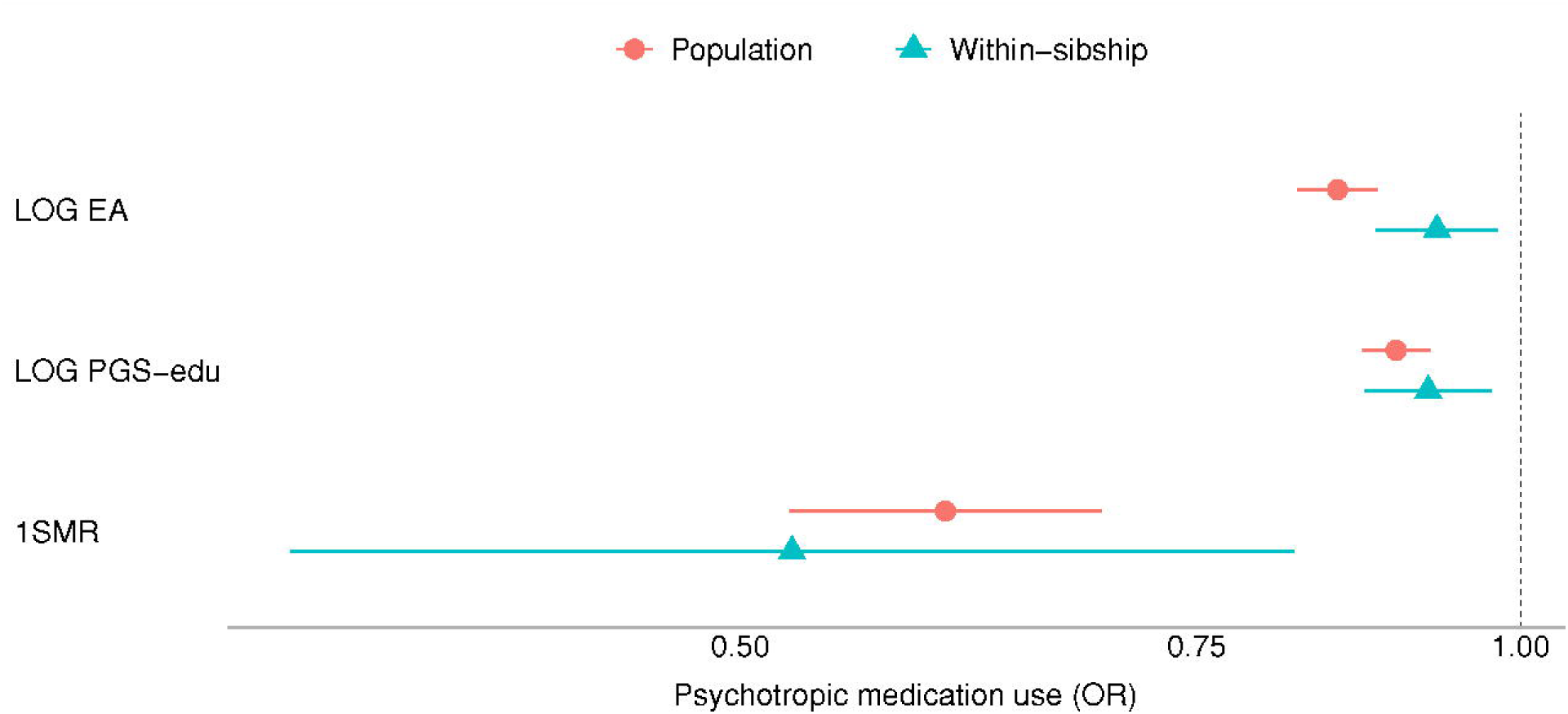
Educational attainment and consumption of psychotropic medication. Log odds changes in consumption of psychotropic medication and its 95% confidence interval per SD increase in years of education are shown. Estimated associations are displayed for logistic regression (LOG) and Mendelian randomization models. *Abbreviations: SD, standard deviation unit; LOG EA, logistic regression model with educational attainment as exposure; LOG PGS-edu, logistic regression model with the educational attainment polygenic score as exposure; 1SMR, one-sample Mendelian randomization*.

Two-sample MR analyses were performed for depression and neuroticism and in both cases, results were consistent with those from one-sample MR and likewise suggest a protective role of education (eTable9). Each SD increase in EA caused a −0.22 [−0.25, −0.18] and −0.21 [−0.25, −0.17] SD decrease in the scores of depression and neuroticism. Weighted median, weighted mode and MR-Egger estimators were consistent with IVW. In both cases, the directionality test indicated that the causal direction is likely to be correct, and that horizontal pleiotropy is unlikely (eTable 9).

Within-sibship two-sample MR estimates suggested an attenuation of both associations, which were however less precise, and overlapped with those of the population-based MR and the null.

## Discussion

The objective of the present study was to assess to what extent the relationships between EA and anxiety, depression, neuroticism and consumption of psychotropic medication are explained by family and/or demographic factors in adulthood. Regression and population-based MR analyses indicate that higher EA is associated with lower levels of anxiety, neuroticism, and consumption of psychotropic medication, but not depression. The protective effect of EA attenuated after accounting for family and/or demographic factors. The observed attenuation was however modest, which is indicative of a causal association between EA and the studied mental health outcomes.

Various potential mechanisms that could explain a causal relationship between EA and the propensity to develop mental health symptoms have been described, and include availability of resources, cognition, knowledge and social integration ^35^. Higher educated people are more likely to have higher income than less educated people, and this difference in earning can affect health by providing easier access to health care services, better nutrition and opportunities for being physical active. Similarly, EA can influence cognition, which can affect health outcomes by leading people to adopt healthier lifestyles and avoiding risk factors associated with mental health conditions such as stress.

However, as family is a driving force of adult health, the existence of other pathways than those in adulthood should be considered ^9–12,36^. Such pathways might involve environment-mediated genetic factors during early life (specially pertained to parents’ characteristics such as genetic-nurturing), similar but reasonably less strong genetic nurturing effect might be expected from the interaction of the individual and other relatives (dynastic-effects) ^17^.

EA might also associate with mental health outcomes because of parental cross-trait assortative-mating on the exposure and outcome or on variables genetically correlated with them (e.g., highly educated individuals are more likely to select partners with lower neuroticism symptomatology) ^20,37^. In some situations parental single-trait assortative-mating on a phenotype genetically correlated with both educational level and the mental health outcome might also explain their association (e.g., partner selection based on cognitive ability that is correlated with EA and the mental health outcome) ^20,38^. Nonetheless, with the present design we are unable to estimate the relative influence of dynastic effects isolated from that of assortative mating or that of population structure.

Since we observed that the level of change was dissimilar among the examined MHC, it is possible that the contribution of indirect genetic effects may vary depending on the outcome of interest and its clinical severity. Accordingly, MHC whose aetiology cannot be fully attributed to direct genetic effects may be influenced by genetic nurturing to a higher extent in comparison to others with a stronger genetic basis. For example, a less pronounced change may be expected for the association between EA and attention-deficit/hyperactivity disorder or schizophrenia, since both seem to rely strongly on direct genetic effects ^39,40^.

### Strengths and limitations

A major strength of our work is that we applied a robust method to evaluate the impact dynastic effects, assortative mating, and population structure may have on the association between EA and the risk of suffering a MHC ^16,33^. As genetic variants are randomly assigned at conception the design is generally likely to avoid bias due to confounding or reverse causality. Moreover, the application of a within-family design has allowed us to overcome the limitations of population-based MR such as non-compliance with the independence criterion ^16^. Importantly, our results were replicated in two different cohorts and across different analyses: Results of observational and MR models were consistent in both cohorts. Nevertheless, our approach has limitations that must be considered when interpreting our findings.

For highly polygenic phenotypes such as EA, limitations arise from the uncertainty regarding the compliance of core assumptions including relevance and exclusion criteria ^36^. Even though first-stage F-statistic indicated our analyses did not suffer from weak instrument bias, our results might still be underpowered to detect an effect of interest, as the PGS-edu only explained nearly 2% of the variation on EA. Similarly, we found little evidence of horizontal pleiotropy.

Importantly, the phenotypical expression of the genetic architecture of EA is contingent on social, historical, and cultural contexts, which might limit the informativeness of the genetic instruments in real scenarios ^36^. EA in the context of MR can be thought of as propensity to achieve a given level of education, which is environmental-dependent, and hence the impact of EA on MHC may vary depending on the environmental context ^36^. Moreover, MR may not be well suited to inform either novel or actionable policy recommendations, as it does not provide much insight on how or why EA is associated to any of the specific MHC we analysed ^36^.

Our within-family analyses had limited power, and could not rule out a causal effect EA on none of the analysed MHC in adulthood. Additionally, as MR relies on available GWAS data (e.g., to calculate PGS-edu) our study suffers from lack of representativeness of populations from other geographical contexts and ancestry which might limit the generalizability of our findings. Our results may be affected by selection bias, as participants of large studies such as UK Biobank were non-randomly sampled ^41^. However, simulations have shown that in the context of MR the impact of selection bias is likely to be less than that of other biases such as pleiotropy or population stratification ^42^.

## Conclusions

Our findings suggest that previously observed associations between EA and MHC in adulthood may to some extent originate and be explained by family-related factors (dynastic effects) and/or assortative mating, and population structure. However, overall all estimates remain robust, indicative of a causal protective effect of EA on the risk of suffering MHC. Future larger family based studies are needed to provide more precise evidence about these effects.

## Supporting information

eMethods

eTables

eTable2

## Data Availability

Researchers associated with Norwegian research institutes can apply for the use of HUNT material: data and samples - given approval by a Regional Committee for Medical and Health Research Ethics. Researchers from other countries can also apply in cooperation with a Norwegian Principle Investigator. Information for data access can be found at https://www.ntnu.edu/hunt/data. UK Biobank individual-level participant data are available via enquiry to. All GWAS summary statistics used in the present manuscript are publicly available and can be download from https://gwas.mrcieu.ac.uk/ and https://thessgac.com/papers/.

## Acknowledgments

The Trøndelag Health Study (HUNT) is a collaboration between HUNT Research Center (Faculty of Medicine and Health Sciences, NTNU, Norwegian University of Science and Technology), Trøndelag County Council, Central Norway Regional Health Authority, and the Norwegian Institute of Public Health. The genotyping in HUNT was financed by the National Institutes of Health; University of Michigan; the Research Council of Norway; the Liaison Committee for Education, Research and Innovation in Central Norway; and the Joint Research Committee between St Olavs hospital and the Faculty of Medicine and Health Sciences, NTNU. We also like to thank to Dr. David Carslake for his his valuable comments.

## Information on author access to data

MFVV and LB had full access to all of the data in the study and take responsibility for the integrity of the data and the accuracy of the data analysis.

## Authors’ contributions

Conception and design: MFVV, LB, BMB and ØEN. Analysis: MFVV and LB. Interpretation of data for the manuscript: MFVV, LB, PRJ, MT, GDS, NMD, BMB and ØEN. Drafting of the manuscript: MFVV and LB. Critical review of the manuscript for important intellectual content: MFVV, LB, PRJ, MT, GDS, NMD, BMB and ØEN. Final approval of the version to be published: MFVV, LB, PRJ, MT, GDS, NMD, BMB and ØEN. Agreement to be accountable for all aspects of the work: MFVV, LB, PRJ, MT, GDS, NMD, BMB and ØEN.

