## Supplementary material for "Educational attainment and mental health conditions: a within-sibship Mendelian randomization study": eMethods

**Cohorts**

*The Trøndelag Health Study (HUNT)*

HUNT is a population-based cohort study that is held in the Trøndelag County in Norway and started in 1984, which is led by the Norwegian University of Science and Technology (NTNU) (1). All adult residents in the Trøndelag County have been invited to participate to each survey: HUNT1 (1983 – 1986), HUNT2 (1995 – 1996), HUNT3 (2006 – 2008) and HUNT4 (2017 – 2019) (1). In the present study we used data from participants of HUNT2 (n = 56 374) and HUNT3 (n = 38 278) who had been genotyped. From those, we selected all individuals that were > 30 years of age when they participated in the survey and had at least one sibling. The final sample from HUNT2 included 26 770 siblings who belonged to 10 428 sibships. The final sample from HUNT3 included 16 718 siblings who belonged to 7010 sibships. Siblings were identified using KING software, with sibling-pairs identified based on the following criteria: i) kinship coefficient between 0.177 and 0.355, ii) the proportion of the genomes that share two alleles IBD > 0.08, and iii) the proportion of the genome that share zero alleles IBD > 0.04. Sibships of two or more siblings were constructed based on the identified sibling-pairs (2).

*UK Biobank*

The UK Biobank is a prospective cohort study that investigates the contribution of genetic and environmental exposures to the development of disease, which began in 2006 in the United Kingdom (UK). UK Biobank is following nearly 500 000, 40 to 69 years old participants from across the UK who volunteered to be part of the study and provided consent for follow-up through linkage to their health records (3). We included all participants who had been genotyped, that is n = 488 377. After restricting the sample to sibships with two or more individuals, our analysis sample included 40,734 individuals from 19,773 sibships. Siblings were identified in a previous study using the UK Biobank derived

estimates of pairwise identical by state (IBS) kinships and the proportion of unshared loci (IBS0) (4). Briefly, sibling-pairs were identified based on the following criteria: i) sibling pairs should have IBS0 > 0 and should have an expected

IBS kinship of 0.5 with a standard deviation of 0.038, ii) cluster of pairwise relationships that labelled the individuals as sibling pairs if they fell within the following bounds (IBS: > 0.5-21*IBS0, < 0.7) and (IBS0: >0.001, <0.008).

*Ethics approval*

The study protocol was approved by the Regional Committees for Medical Research Ethics South East (REK 2017/2479) and Mid-Norway (REK 2015/1197). All participants signed informed consent for participation and the use of data in research. UK Biobank obtained ethics approval from the North West Multi-centre Research Ethics Committee and obtained informed consent from all study participants.

**Genotyping**

*HUNT*

There were 70 517 genotyped individuals who participated in either HUNT2 or HUNT3 (5). Standard HumanCoreExome arrays (HumanCoreExome12 v1.0 and v1.1) were used to genotype 12 864 samples and a customised HumanCoreExome array (UM HUNT Biobank v1.0) for the remainder. Quality control was performed separately for genotype data from different arrays. The call rate of genotyped samples was > 99%. Imputation was performed on samples of recent European ancestry using Minimac3 (v2.0.1) from a merged reference panel constructed from the Haplotype Reference Consortium panel (release version 1.1) and a merged local reference panel based on 2201 whole-genome sequenced HUNT participants.

*UK Biobank*

Information on genotyping and quality control has been described elsewhere (3). Applied Biosystems UK BiLEVE Axiom Arrays by Affymetrix (now part of Thermo Fisher Scientific) were used to genotype a subset of 49 950, and the closely related Applied Biosystems UK Biobank Axiom Array for the remainder. Technical details of Affymetrix’s laboratory process are available in (6), and details of the genotyping calling routine specific to the UK Biobank project are available in (7). Imputation was performed using Impute4 software and UK10K reference panels. We used genomic data from the release version 3 and the Human GRCh37 (hg19) human reference genome for the analyses (3).

**Genetic instrument**

*HUNT*

From the 3952 single nucleotide polymorphisms (SNPs) reported in the GWAS, we included those that were well imputed (r2 >= 0.80 and MAF <= 0.001) in the target population. PLINK was used to estimate allele dosages and weighted PGS, applying the PLINK function: -‑score. Weights for the polygenic score (PGS) were extracted from (8). For HUNT participants, the PGS-edu consisted of a weighted sum of of the allele dosages of 3781 SNPs. For UKB participants, the PGS-edu consisted of a weighted sum of of the allele dosages of 3758 SNPs (see eTable 2 for a list of the SNPs that were included in both PGS).

**Phenotypes**

*Educational attainment*

In UK Biobank number of years of education was assigned according to the methodology described by Okbay et. al., which is based on The International Standard Classification of Education (ISCED) mapping 1997 (8,11). EA corresponded to data-field 6138. EA was not measured in HUNT3 and therefore for participants of HUNT3 we used EA reported in HUNT4 and if it was missing EA from HUNT2. Either case a number of years of education was assigned according to ISCED mapping 1997 (11). See eTable 2 to check the specific number of years of education that was assigned to the answers to the questions: w*hat is your highest level of education?* (HUNT participants) and *which of the following qualifications do you have? (You can select more than one)?* (UK Biobank participants)?

*Anxiety and depression symptomatology*

In HUNT symptoms of anxiety and depression were assessed by the Hospital Anxiety and Depression Scale (HADS), which is a self-assessment 14 item scale with a score that ranges from 0 to 42 (12). HADS derives from summing the responses of two subscales, one for depression and one for anxiety, each with a score that ranges from 0 to 21. The higher the score the higher the level of anxiety or depression a person is experiencing. We analysed separately the score for anxiety (HADS-anx) and that for depression (HADS-dep). A recommended cut point of 10 over 21 for each of the subscales was used to identify cases of anxiety and depression (12).

In UK Biobank symptoms of anxiety and depression were evaluated by the 7-item Generalized Anxiety Disorder Scale (GAD‐7) and 9-item Patient Health Questionnaire (PHQ-9), respectively. Both, were part of a self-completion mental health questionnaire that was responded by a total of 157 366 individuals aged 42 to 81 years old (13). GAD-7 consists of seven items that score DSM-IV criteria upon which the diagnosis of generalized anxiety disorder is based. The GAD-7 score derives from summing the responses of the seven items, each scored from 0 (not al all) to 3 (nearly every day), and ranges from 0 to 21 (14). The higher the score the higher the level of anxiety symptomatology a person is experiencing. UK Biobank fields used to calculate the GAD-7 score were 20505, 20506, 20509, 20512, 20515, 20516, and 20520.

The PHQ-9 consists of nine items that score DSM-IV criteria upon which the diagnosis of depressive disorders is based. The PHQ-9 score derives from summing the responses of the nine items, each scored from 0 (not al all) to 3 (nearly every day), and ranges from 0 to 27 (15). The higher the score the higher the level of depressive symptomatology a person is experiencing. UK Biobank fields used to calculate the PHQ-9 score were 20507, 20508, 20510, 20511, 20513, 20514, 20517, 20518, and 20519. A recommended cut point of 10 over 21 or 27 for each of the scales was used to identify cases of anxiety and depression.

*Neuroticism*

Neuroticism was assessed the Eysenck Personality Questionnaire, which is a self-assessment scale designed to evaluate three personality dimensions: extraversion, neuroticism, and psychoticism (16). In HUNT a shortened version of the questionnaire (six items) was applied, and two dimensions of personality were measured: extraversion and neuroticism, each one assessed by six questions with two possible answers yes (scored 1) or no (scored 0). For the present study we selected questions related to neuroticism, so the score of neuroticism varied from 0 to 6. The higher the score the higher the level of neuroticism a person is experiencing.

In UK Biobank the 12 item Eysenck Personality Questionnaire was applied. Each of the items corresponded to data-fields 1920, 1930, 1940, 1950, 1960, 1970, 1980, 1990, 2000, 2010, 2020, and 2030 (16). The externally derived score, which corresponded to data-field 20127, derives from summing the responses of the 12 items, each scored 0 (no) or 1 (yes), and ranges from 0 to 12. The higher the score the higher the level of neuroticism a person is experiencing.

*Consumption of psychotropic medication*

In HUNT participants were asked whether they have ever taken daily medication. If the answer was “yes”, they were then asked which medication and for how long (months) the medication was consumed. The number of months of consumption (from 0 to 12) of three groups of psychotropic drugs was evaluated: antidepressants, sleep medication and sedatives. For the present analysis we summed the number of months of consumption of the aforementioned drugs, and then identified two groups of individuals: people who reported no usage of psychotropic medication (0 months) and people who reported usage of psychotropic medication (> 0 months). Consumption of psychotropic medication was self-reported only in HUNT2 (n = 52 806).

In UK Biobank medication status was obtained from data-field 20003 which corresponded to the names of regular prescription medications (47 drug codes) that participants reported having currently taking during a verbal interview. From the 47 drug codes we selected those usually prescribed to treat MHC and mapped them to four groups: antidepressants, anxiolytics, mood stabilisers, and antipsychotics according to a list retrieved from (17). We then counted the number of any MHC related medication by person and identified two groups of individuals: people who reported no usage of psychotropic medication (0 MHC related medication) and people who reported usage of psychotropic medication (> 0 MHC related medication). We dropped cases when a person had missing information on the field 137 that corresponds to number of medications taken.

*Imputation*

We used predictive mean matching to impute the continuous variables, and polytomous logistic regression to impute categorical variables. We generated 20 imputed datasets with 20 iterations for each imputation. Trace plots were inspected to determine whether the algorithm converged. Generation of imputed data sets, trace plots, and pooling of analyses was performed using the R package mice (3.15.0) (18).

*Sibling analyses*

Models including family fixed effects included a dummy variable for each sibship, which allowed to account for any difference between families due to indirect genetic effects (i.e. genetic nurturing) or bias due to assortative mating (4). Fixed effect models for symptoms of anxiety, depression and neuroticism (continuos outcomes) were specified following the recommendations from the user manual of the “fixest” package, which also allows to perform instrumental variables analysis, but only for numerical outcomes (19). The models in the case were specified as follows:

**1. OLS + clustered standard errors by sibship ID**

feols (outcome ~ exposure + covariable 1 + covariable 2 …, ~ sibship ID, data)

**2. OLS + clustered standard errors by sibship ID + family fixed effects**

feols (outcome ~ exposure + covariable 1 + covariable 2 … | sibship ID, ~ sibship ID, data)

**3. Mendelian randomization + clustered standard errors by sibship ID**

feols (outcome ~ exposure + covariable 1 + covariable 2 … | exposure ~ genetic instrument, ~ sibship ID, data)

**4. Mendelian randomization + clustered standard errors by sibship ID + family fixed effects**

feols (outcome ~ exposure + covariable 1 + covariable 2 … | sibships ID | exposure ~ genetic instrument, ~ sibship ID, data)

Consumption of psychotropic medication (dichotomous outcome) was modelled instead using the feglm function of the “fixest” package. Difference between families due to indirect genetic effects (i.e. genetic nurturing) or bias due to assortative mating in this case was accounted for by applying the sibling difference method, following the recommendations from (4). The approach is based on the estimation of i) the mean of the exposure among the siblings (family mean) and ii) the estimation of the exposure centered on the sibling mean (exposure centered), which are included in the model as dummy variables. The centered mean corresponds to the difference between the exposure and the family mean. The models in the case were specified as follows:

**5. LOG + clustered standard errors by sibship ID**

feglm (outcome ~ exposure + covariable 1 + covariable 2 …, ~ sibship ID, family = binomial (), data)

**6. LOG + clustered standard errors by sibship ID + family fixed effects**

feglm (outcome ~ exposure + covariable 1 + covariable 2 … + family mean + exposure centered, ~ sibship ID, family = binomial (), data)

Since it was not possible to run instrumental variables analysis for Consumption of psychotropic medication (dichotomous outcome) using the “fixest” package we apply the two step approach. The models in the case were specified as follows:

**7. Mendelian randomization + clustered standard errors by sibship ID**

First stage

feols (exposure ~ genetic instrument + covariable 1 + covariable 2 …, ~ sibship ID, data)

Second stage

feglm (outcome ~ fitted values for exposure from the first stage + covariable 1 + covariable 2 …, ~ sibship ID, family = binomial (), data)

**8. Mendelian randomization + clustered standard errors by sibship ID + family fixed effects**

First stage

feols (exposure centered ~ genetic instrument centered + covariable 1 + covariable 2 …, ~ sibship ID, data)

Second stage

feglm (outcome ~ fitted values for exposure centered from the first stage + covariable 1 + covariable 2 …, ~ sibship ID, family = binomial (), data)
