## Supplementary material for "Educational attainment and mental health conditions: a within-sibship Mendelian randomization study": eTables

**eTable 1. Description of GWAS used in one and two-sample MR analyses.**

| **Variable** | **PMID** | **Author (year)** | **Downloaded from** | **Consortium** | **Population** | **Sample size** | **Sex female** |
| --- | --- | --- | --- | --- | --- | --- | --- |
| Years of education (SD) | 35361970 | Okbay, A., et al. (2022) (1) | https://thessgac.com/papers/ | SSGAC | European/population | 3 037 499 | Mixed* |
| Years of education (SD) | 35534559 | Howe, L.J., et al. (2022) (2) | https://gwas.mrcieu.ac.uk/datasets/ieu-b-4835/ | The Within Family Consortium | European/sibling | 54 986 | Mixed* |
| Depressive symptoms (SD) | **27089181** | Okbay, A., et al. (2016) (3) | https://gwas.mrcieu.ac.uk/datasets/ieu-a-1000/ | SSGAC | European/population | 161 460 | Mixed* |
| Depressive symptoms (SD) | 35534559 | Howe, L.J., et al. (2022) (2) | https://gwas.mrcieu.ac.uk/datasets/ieu-b-4839/ | The Within Family Consortium | European/sibling | 16 782 | Mixed* |
| Neuroticism (SD) | **27089181** | Okbay, A., et al. (2016) (3) | https://gwas.mrcieu.ac.uk/datasets/ieu-a-1007/ | SSGAC | European/population | 170 910 | Mixed* |
| Neuroticism (SD) | 35534559 | Howe, L.J., et al. (2022) (2) | https://gwas.mrcieu.ac.uk/datasets/ieu-b-4847/ | The Within Family Consortium | European/sibling | 28 130 | Mixed* |

***Abbreviations and symbology:*** *SSGAG, Social Science Genetic Association Consortium; *, information on the sex proportion for the whole sample was not reported.*

**eTable 2. Sample sizes.**

|  | **HUNT** | | **UKB** | | **Total** | |
| --- | --- | --- | --- | --- | --- | --- |
| **Outcome** | **Siblings** | **Sibships** | **Siblings** | **Sibships** | **Siblings** | **Sibships** |
| Anxiety | 26 770 | 10 428 | 4863 | 2404 | 31 633 | 12 832 |
| Depression | 26 770 | 10 428 | 4863 | 2404 | 31 633 | 12 832 |
| Neuroticism | 16 718 | 7010 | 23 852 | 11 655 | 40 570 | 18 665 |
| Psych. med. | 26 770 | 10 428 | 35 118 | 17 079 | 61 888 | 27 507 |

**Abbreviations and symbology:** UKB, UK Biobank; Psych. med., consumption of psychotropic medication.

**eTable 3. List of SNPs included in the education polygenic score (PGS-edu)**

**eTable 4. Cohort level EA measures**.

| **HUNT** | |
| --- | --- |
| *What is your highest level of education?*  *HUNT 2*  1) Primary school 7-10 years, continuation school, folk high school  2) High school, intermediate school, vocational school, 1-2 years high school  3) University qualifying examination, junior college, A levels  4) University or other post-secondary education, less than 4 years  5) University/college, 4 years or more  *HUNT 4*  1) 9-10 years compulsory primary and lower secondary school  2) One or two years of academic or vocational school  3) 3 years of academic or vocational school  4) 3-4 years vocational school/apprentice (upper secondary/sixth form college)  5) College or university, less than four years  6) College or university, four years or more | Assigned years of education  *HUNT 2*  1) 10 2) 13 3) 15 4) 16 5) 21  *HUNT 4*  1) 10 2) 12 3) 13 4) 15 5) 16  6) 21 |
| **UK Biobank** | |
| *Which of the following qualifications do you have? (You can select more than one)?*  1) College or University degree 2) A levels/AS levels or equivalent 3) O levels/GCSEs or equivalent  4) CSEs or equivalent 5) NVQ or HND or HNC or equivalent 6) Other prof. qual. eg: nursing, teaching  7) None of the above  8) Prefer not to answer | Assigned years of education  1) 20 2) 13 3) 10 4) 10 5) 14 6) 15 7) 7 8) Excluded  (Highest category assigned to respondents who selected multiple options) |

**eTable 5. Summary of model statistics for each of the meta-analyses.**

| **Model** | **Model statistics** | **Model** |
| --- | --- | --- |
| Anxiety |  |  |
| OLS EA | Q(df = 1) = 7.81, *p = 0.005*, I^2^ = 87.20% | Random-effects |
| OLS EA + FE | Q(df = 1) = 0.84, *p = 0.360*, I^2^ = 00.00% | Fixed-effects |
| OLS PGS-edu | Q(df = 1) = 1.18, *p = 0.277*, I^2^ = 15.38% | Fixed-effects |
| OLS PGS-edu + FE | Q(df = 1) = 3.39, *p = 0.066*, I^2^ = 70.50% | Fixed-effects |
| 1SMR | Q(df = 1) = 0.12, *p = 0.727*, I^2^ = 00.00% | Fixed-effects |
| 1SMR + FE | Q(df = 1) = 3.19, *p = 0.074*, I^2^ = 68.68% | Fixed-effects |
| Depression |  |  |
| OLS EA | Q(df = 1) = 2.40, *p = 0.121*, I^2^ = 58.41% | Fixed-effects |
| OLS EA + FE | Q(df = 1) = 2.30, *p = 0.129*, I^2^ = 56.56% | Fixed-effects |
| OLS PGS-edu | Q(df = 1) = 19.27, *p < 0.001*, I^2^ = 94.81% | Random-effects |
| OLS PGS-edu + FE | Q(df = 1) = 10.70, *p = 0.001*, I^2^ = 90.65% | Random-effects |
| 1SMR | Q(df = 1) = 14.51, *p < 0.001*, I^2^ = 93.11% | Random-effects |
| 1SMR + FE | Q(df = 1) = 7.18, *p = 0.007*, I^2^ = 86.07% | Random-effects |
| Neuroticism |  |  |
| OLS EA | Q(df = 1) = 9.39, *p = 0.002*, I^2^ = 89.35% | Random-effects |
| OLS EA + FE | Q(df = 1) = 3.43, *p = 0.064*, I^2^ = 70.86% | Random-effects |
| OLS PGS-edu | Q(df = 1) = 5.90, *p = 0.015*, I^2^ = 83.05% | Random-effects |
| OLS PGS-edu + FE | Q(df = 1) = 1.23, *p = 0.2672*, I^2^ = 18.76% | Fixed-effects |
| 1SMR | Q(df = 1) = 17.73, *p < 0.001*, I^2^ = 94.36% | Random-effects |
| 1SMR + FE | Q(df = 1) = 1.45, *p = 0.229*, I^2^ = 30.81% | Fixed-effects |
| Psych. med. |  |  |
| LOG EA | Q(df = 1) = 0.07, *p = 0.789*, I^2^ = 00.00% | Fixed-effects |
| LOG EA + FE | Q(df = 1) = 0.75, *p = 0.387*, I^2^ = 00.00% | Fixed-effects |
| LOG PGS-edu | Q(df = 1) = 0.01, *p = 0.911*, I^2^ = 00.00% | Fixed-effects |
| LOG PGS-edu + FE | Q(df = 1) = 0.24, *p = 0.621*, I^2^ = 00.00% | Fixed-effects |
| 1SMR | Q(df = 1) = 0.59, *p = 0.443*, I^2^ = 00.00% | Fixed-effects |
| 1SMR + FE | Q(df = 1) = 0.30, *p = 0.584*, I^2^ = 00.00% | Fixed-effects |

**Abbreviations and symbology:** OLS, ordinary least squares regression; EA, educational attainment; FE, within-sibship adjustment; PGS-edu, educational attainment polygenic score; 1SMR, one-sample Mendelian randomization; Psych. med., psychotropic medication usage; LOG, logistic regression.

**eTable 6. Missing data in HUNT**.

| **Variable** | **HUNT2** | | |  | **HUNT3** | | |
| --- | --- | --- | --- | --- | --- | --- | --- |
|  | **n** | **Total** | **%** |  | **n** | **Total** | **%** |
| Education attainment | 1398 | 26 770 | 5.22 |  | 1216 | 16 718 | 7.27 |
| Sleep medication usage | 4448 | 26 770 | 16.62 |  | - | - | - |
| Sedatives usage | 4446 | 26 770 | 16.61 |  | - | - | - |
| Antidepressive medication usage | 4435 | 26 770 | 16.57 |  | - | - | - |
| **HADS** |  |  |  |  |  |  |  |
| Nervous | 3154 | 26 770 | 11.78 |  | - | - | - |
| Catastrophe | 2052 | 26 770 | 7.67 |  | - | - | - |
| Worry | 1824 | 26 770 | 6.81 |  | - | - | - |
| Slow | 1677 | 26 770 | 6.26 |  | - | - | - |
| Restless 1 | 1462 | 26 770 | 5.46 |  | - | - | - |
| Panic | 1390 | 26 770 | 5.19 |  | - | - | - |
| Restless 2 | 1343 | 26 770 | 5.02 |  | - | - | - |
| Enjoy | 1132 | 26 770 | 4.23 |  | - | - | - |
| Optimistic | 1136 | 26 770 | 4.24 |  | - | - | - |
| Laugh | 1096 | 26 770 | 4.09 |  | - | - | - |
| Look | 1040 | 26 770 | 3.88 |  | - | - | - |
| Happy | 949 | 26 770 | 3.55 |  | - | - | - |
| Relaxed | 947 | 26 770 | 3.54 |  | - | - | - |
| Media | 909 | 26 770 | 3.40 |  | - | - | - |
| **EPQ** |  |  |  |  |  |  |  |
| Humiliation | - | - | - |  | 826 | 16 718 | 4.94 |
| Tired | - | - | - |  | 785 | 16 718 | 4.70 |
| Indifferent | - | - | - |  | 782 | 16 718 | 4.68 |
| Nervous | - | - | - |  | 725 | 16 718 | 4.34 |
| Worry | - | - | - |  | 713 | 16 718 | 4.26 |
| Catastrophe | - | - | - |  | 675 | 16 718 | 4.04 |

**Abbreviations and symbology:** HADS, Hospital Anxiety and Depression Scale; EPQ, Eysenck Personality Questionnaire; GAD-7, 7-item Generalized Anxiety Disorder Scale; PHQ-9, 9-item Patient Health Questionnaire; *, UK Biobank field; N/A, no missing information.

**eTable 7. Association between the education polygenic score (PGS-edu) and the outcomes.**

**eTable 8. Summary of the one-sample MR model statistics.** *Abbreviations and symbology: Stat, test statistic; p, p value; df., degrees of freedom; -FE, within-sibship adjustment; Psych. med., psychotropic medication usage.*

|  | **HUNT** | | | | | | **UK Biobank** | | | | | |
| --- | --- | --- | --- | --- | --- | --- | --- | --- | --- | --- | --- | --- |
|  | **F-test (1st stage)** | | | **Wu-Hausman** | | | **F-test (1st stage)** | | | **Wu-Hausman** | | |
|  | **Stat.** | ***p*** | **df.** | **Stat.** | ***p*** | **df** | **Stat.** | ***p*** | **df.** | **Stat.** | ***p*** | **df** |
| **Anxiety** |  |  |  |  |  |  |  |  |  |  |  |  |
| 1SMR | 1162.79 | 6.89x10^-246^ | 26755 | 17.18 | 4.52x10^-5^ | 26754 | 318.00 | 2.20 x 10^-16^ | 4848 | 12.40 | 4.36 x 10^-4^ | 4847 |
| 1SMR + FE | 364.08 | 5.48x10^-77^ | 26755 | 0.30 | 0.611 | 16327 | 34.10 | 5.64 x 10^-9^ | 4846 | 5.15 | 0.023 | 2444 |
| **Depression** |  |  |  |  |  |  |  |  |  |  |  |  |
| 1SMR | 1162.79 | 6.89x10^-246^ | 26755 | 0.40 | 0.538 | 26754 | 318.00 | 2.20 x 10^-16^ | 4848 | 25.20 | 5.27 x 10^-7^ | 4847 |
| 1SMR + FE | 364.08 | 5.48x10^-77^ | 26755 | 0.08 | 0.806 | 16327 | 34.10 | 5.64 x 10^-9^ | 4846 | 12.50 | 4.22 x 10^-4^ | 2444 |
| **Neuroticism** |  |  |  |  |  |  |  |  |  |  |  |  |
| 1SMR | 573.53 | 3.40x10^-121^ | 16703 | 54.20 | 5.72x10^-13^ | 16702 | 1520.00 | 2.20 x 10^-16^ | 23 837 | 22.70 | 1.86 x 10^-6^ | 23 836 |
| 1SMR + FE | 206.88 | 1.76x10^-44^ | 16703 | 9.57 | 0.003 | 9693 | 301.20 | 2.20 x 10^-16^ | 23 837 | 4.21 | 0.040 | 12 182 |
| **Psych. med.** |  |  |  |  |  |  |  |  |  |  |  |  |
| 1SMR | 1162.79 | 6.89x10^-246^ | 26755 | 13.78 | 3.27x10^-4^ | 26754 | 2230.30 | 2.20 x 10^-16^ | 35 103 | 15.70 | 7.63 x 10^-5^ | 35 102 |
| 1SMR + FE | 364.08 | 5.48x10^-77^ | 26755 | 1.95 | 0.197 | 16327 | 373.20 | 2.20 x 10^-16^ | 35 103 | 4.89 | 0.027 | 18 024 |

**eTable 9. Summary of two-sample MR estimators.** Two-sample MR analyses were performed using summary statistics from the GWAS described in Supplementary table 1. Results are shown for two genetic instruments for EA. *Abbreviations and symbology: SNP, single nucleotide polymorphism; n, number; SE, standard error; p, p value; LCI, lower confidence interval; UCI, upper confidence interval; W, weighted; S, simple; IVW, inverse variance weighted; -, not available.*

|  | **Estimators** | **Population** | | | | | | **Within-sibling** | | | | | |
| --- | --- | --- | --- | --- | --- | --- | --- | --- | --- | --- | --- | --- | --- |
|  |  | **SNPs (n)** | **B** | **SE** | ***p*** | **LCI** | **UCI** | **SNPs (n)** | **B** | **SE** | ***p*** | **LCI** | **UCI** |
| **Depressive symptoms (Okbay et al.)**  *MR-Steiger:* r2 exposure: 0.016, r2 outcome: 0.006; *p = 4.30x10^-97^*  *Egger intercept* = 0.000 (SE, 0.001); *p = 0.435* | MR Egger | 560 | -0.18 | 0.06 | 0.002 | -0.28 | -0.07 | - | - | - | - | - | - |
|  | W median | 560 | -0.19 | 0.02 | 2.72x10^-14^ | -0.24 | -0.14 | - | - | - | - | - | - |
|  | IVW | 560 | -0.22 | 0.02 | 5.34x10^-34^ | -0.25 | -0.18 | - | - | - | - | - | - |
|  | S mode | 560 | -0.30 | 0.09 | 0.001 | -0.48 | -0.12 | - | - | - | - | - | - |
|  | W mode | 560 | -0.16 | 0.06 | 0.012 | -0.29 | -0.04 | - | - | - | - | - | - |
| **Depressive symptoms (Howe et al.)** | MR Egger | 489 | -0.18 | 0.12 | 0.116 | -0.41 | 0.04 | 497 | -0.01 | 0.12 | 0.904 | -0.25 | 0.22 |
|  | W median | 489 | -0.16 | 0.05 | 0.001 | -0.25 | -0.06 | 497 | -0.16 | 0.11 | 0.147 | -0.37 | 0.05 |
|  | IVW | 489 | -0.14 | 0.04 | 0.000 | -0.21 | -0.07 | 497 | -0.05 | 0.07 | 0.494 | -0.19 | 0.09 |
|  | S mode | 489 | -0.37 | 0.18 | 0.038 | -0.71 | -0.02 | 497 | -0.07 | 2.93 | 0.981 | -5.80 | 5.67 |
|  | W mode | 489 | -0.35 | 0.16 | 0.020 | -0.65 | -0.06 | 497 | -0.07 | 2.71 | 0.980 | -5.38 | 5.25 |
| **Neuroticism (Okbay et al.)**  *MR-Steiger:* r2 exposure: 0.016, r2 outcome: 0.008; *p = 1.47x10^-63^*  *Egger intercept* = 0.000 (SE, 0.001); *p = 0.650* | MR Egger | 560 | -0,24 | 0.06 | 1.91x10^-04^ | -0.37 | -0.11 | - | - | - | - | - | - |
|  | W median | 560 | -0.20 | 0.03 | 1.05x10^-14^ | -0.25 | -0.15 | - | - | - | - | - | - |
|  | IVW | 560 | -0.21 | 0.02 | 3.95x10^-25^ | -0.25 | -0.17 | - | - | - | - | - | - |
|  | S mode | 560 | -0.22 | 0.10 | 0.028 | -0.42 | -0.02 | - | - | - | - | - | - |
|  | W mode | 560 | -0.22 | 0.08 | 0.003 | -0.37 | -0.07 | - | - | - | - | - | - |
| **Neuroticism (Howe et al.)** | MR Egger | 491 | -0.31 | 0.09 | 5.56x10^-04^ | -0.48 | -0.13 | 499 | -0.08 | 0.09 | 0.381 | -0.25 | 0.10 |
|  | W median | 491 | -0.19 | 0.04 | 1.29x10^-06^ | -0.26 | -0.11 | 499 | -0.04 | 0.08 | 0.608 | -0.21 | 0.12 |
|  | IVW | 491 | -0.18 | 0.03 | 4.17x10^-10^ | -0.23 | -0.12 | 499 | -0.10 | 0.05 | 0.053 | -0.21 | 0.00 |
|  | S mode | 491 | -0.24 | 0.15 | 1.14x10^-01^ | -0.54 | 0.06 | 499 | -0.06 | 2.54 | 0.982 | -5.04 | 4.93 |
|  | W mode | 491 | -0.27 | 0.12 | 3.20x10^-02^ | -0.51 | -0.02 | 499 | -0.06 | 2.56 | 0.982 | -5.06 | 4.95 |
